# AI and Hierarchical clustering techniques for accurate patient stratification

**DOI:** 10.64898/2026.03.13.26348331

**Authors:** Juan G. Diaz Ochoa, Miroslav Puskaric, Natalie Layer, Antje Jensch, Markus Knott, Alexander Krohn

## Abstract

Graph-based methods for data representation and analysis are well suited for encoding both data points and their interrelationships. This approach integrates data and topology, enabling the representation of interrelated information. In this study, we represent patient cohorts as cohort graphs and discuss their application for real-world patient data. We particularly focus on developing methods to cluster patients with similar symptoms and examine how bias parameters (such as sex and age group) influence interlinking within CGs, thereby improving results for accurate patient stratification and personalized decision-making in a clinical context. In particular we illustrate how by considering sex and age groups we can improve the symptom-clustering of a patient population with lung and gastro-intestinal cancer. Finally, we discuss the essential role of high-performance computing (HPC) in upscaling analytical methods for CGs.

## 1. Introduction

Graph-based modeling is an efficient way to encode information since it captures not only data but also the interrelationships between observables and parameters. For this reason, graph-based modeling has gained increasing importance in medicine for representing physiological relationships and biomedical outcomes [1], [2]. A key application of graph-based modeling is the encoding of relevant physiological and biomedical information. Recently, several groups have extended such methods to encode patient cohorts to represent patients’ etiology via a similar approach to social networks. This is done by considering diagnoses or symptoms as comparison criteria to establish similarity and thus interlink patients [5].

In this study, we provide a concise overview of the state of the art in **cohort graph modelling** (**CGM**) and discuss both the advantages and limitations of CGM. We cover methodological aspects, such as the precision of patient embedding into networks, as well as the computational limits of available methods, and highlight how high-performance computing (**HPC**) can play a decisive role in the further development of CGs.

## 2. Related work

In recent years, access to electronic health records and the MIMIC database [6] (which contains a systematic record of patients in intensive care units) has opened the door to graph-based patient representations [7]. While several works rely on MIMIC data for benchmarking [5], the real challenge ahead is the implementation of graph-based methods with access to real-world patient data. One way to implement this type of graph modeling while keeping the data safe is by means of data synthesis from real medical data (**RMD**). For example, for patients with renal insufficiency, this approach has been successfully applied to predict medical procedures based on patients with similar symptoms [8]. However, for more complex patient cohorts, such data synthetization techniques might be insufficient.

For example, oncology patients in the Emergency Department (**ED**) present multiple symptoms. Pure recording of the “Chief Complaint” (e.g., using the Canadian Emergency Department Information System CEDIS [9]) reduces complex cases to single labels and removes clinically important information about symptom combinations (e.g., pain + dyspnea + nausea). But, red flags (e.g., fever + neutropenia signs + nausea) are less easily detected via individual scores, whereas full detection increases the sensitivity of detection. Therefore, in such complex situations, RMD together with advanced techniques for the extraction of structured data (for example, physiological data) and unstructured data (for example, clinical narratives) are required to build accurate data– analysis pipelines.

## 3. Methods

As an example of our methodological approach, patients with similar symptoms were represented via patient networks. The core idea is that, like social networks, two patients are linked if they have similar symptoms. This methodology has been tested not only on benchmarked networks but also on real-world patient data. At Klinikum’s ED in Stuttgart, a total of 10,036 patients with an oncological diagnosis were recorded from 2010–2024 (representing 3.91% of all ED presentations).

For unstructured data extraction, we implemented fine-tuned Bio-Gott-BERT methods to automatically extract symptoms, negations and anatomical references of expressed symptoms from the anamnesis texts of oncological ED patients [10]. Furthermore, we implemented an interpatient comparison, assuming that similar patients have similar symptoms. Gender and age play a relevant role in this stratification, as symptoms are associated with both parameters due to patients’ physiology [11], we weighted the symptom comparison using both parameters instead of using only symptoms as comparison criteria. To this end, we introduced the parameter *g*_*ij*_ [1] defined as:

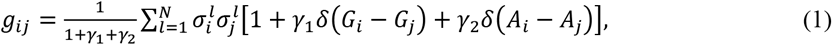

where 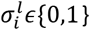 is a binary marker indicating the absence or presence of symptoms *l*ϵ{1, …, *N*} in patient *i* and where *G*_*i*_ and *G*_*j*_ correspond to the sex of patients *i* and *j*, respectively. The function *δ* represents the Kronecker delta, and *γ*_*k*_ and *k* ϵ{1,2} are parameters defining the strength of the bias (set to *γ*_*k*_ = 1, in this study). In previous investigations, we considered only the effect of sex bias on the final clustering [1]. We extended this approach by additionally considering age groups *A*_*i*_ and *A*_*j*_ for patients *i* and *j*, such that patients of similar sex and age exhibit stronger links and form more accurate clusters.

## 4. Results

The results of exemplary hierarchical clustering (Girvan-Newman algorithm) applied to the CG for a cohort of 50 patients are presented in Figure 1 (colored regions).

**Figure 1.**
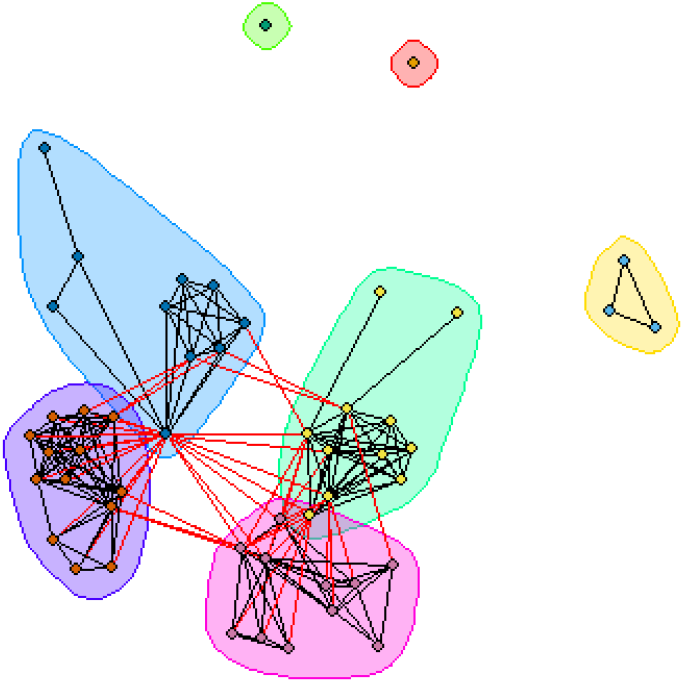
Clustering of a small CG with 50 patients, where each dot represents a single patient. Edges are defined by equation (1). Patients with only one symptom (dots without edges) are, in some cases, clustered in a single cluster (green and yellow clusters).

Hierarchical clustering tends to cluster patients with only one symptom in individual clusters. We added additional bias parameters into the linking function to compensate for this effect. This adjustment effectively reduced the total number of extracted clusters in the CG from 256 to 88 for lung cancer patients and from 325 to 267 for gastrointestinal (GI) cancer patients, as shown in Figure 2. The complexity of lung cancer (a disease often constrained to one organ) is considerably lower than the complexity of GI cancer (which involves multiple organs).

**Figure 2.**
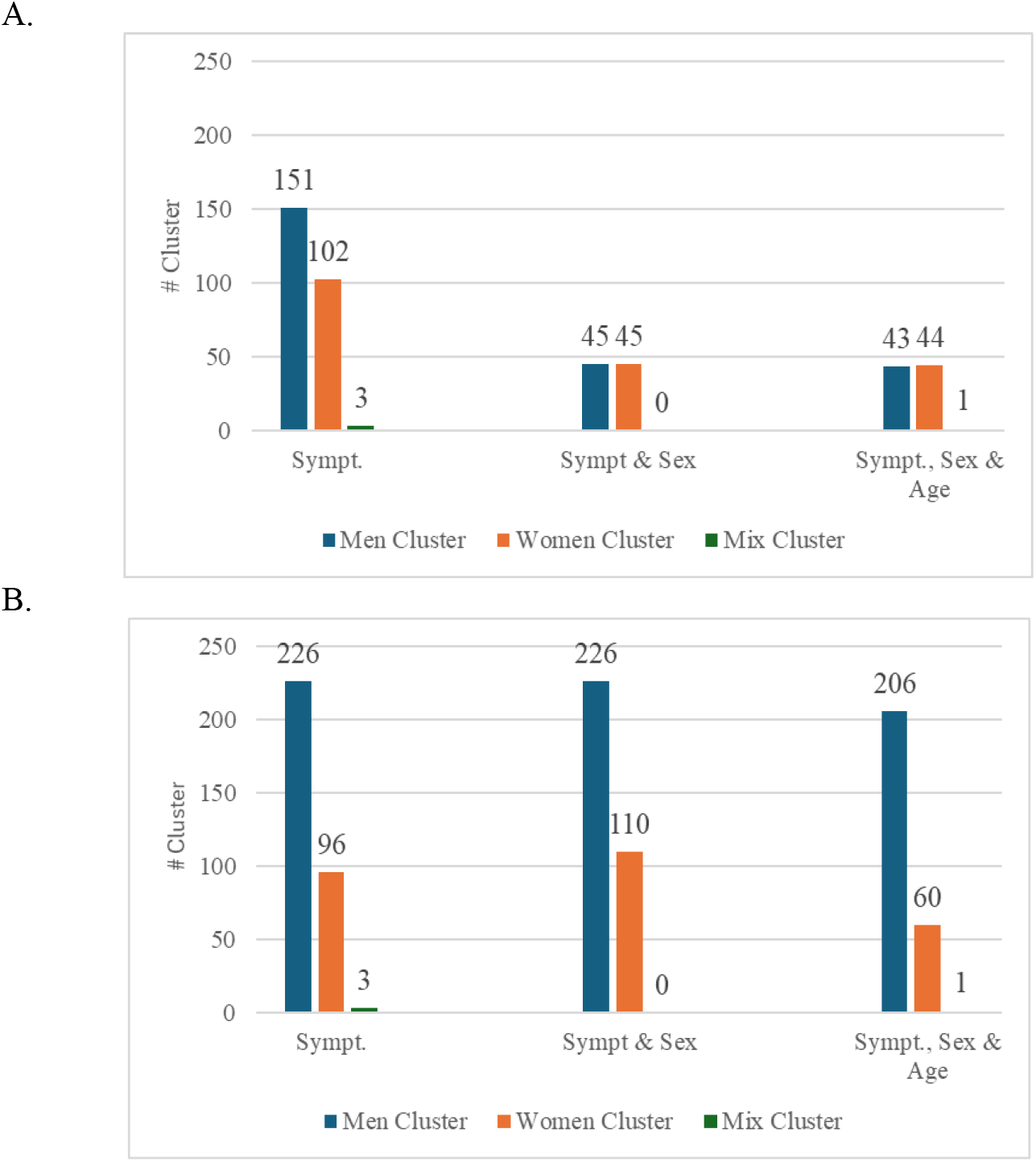
Cluster distribution as a function of model specifications. We analyzed the effect on patient clustering for 500 lung cancer (A) and 500 gastrointestinal cancer (B) patients. The x-axis shows three model configurations: symptom-only, symptom + sex, and symptom + sex + age. The y-axis indicates the number of clusters identified in each configuration. Blue bars cluster only men, orange bars cluster only women, and green bars are mixed clusters.

While modular-clustering methods (such as the Louvain or K-Means algorithm) are computationally efficient, they are not capable to stratify the symptom distribution. Furthermore, the prior probability of the clusters is unknown, impairing the application of the K-Means method. On the other hand, the high frequency of patients showing only one symptom distorts the clustering method, since multiple clusters possess only one patient with one symptom, which is not statistically significant. Incorporating meaningful bias parameters from a clinical perspective, like patient’s age and sex, mitigates this effect and influence the stability of hierarchical clustering. This is in turn helpful for clinical applications like a more precise correlation between symptom groups and diagnoses, for instance.

## 5. Discussion

One limitation of this study is that clinical meaningfulness of the results requires further validation through prospective studies. Furthermore, efficient clustering is required for larger patient populations, but scaling the methodology to enable a meaningful clinical application remains a challenge. Hierarchical clustering has a complexity of 𝒪(*n*^2^), which implies that the computational size increases in a prohibitive way when the network’s size (number of individuals in the cohort *n*) increases.

High-performance computing (HPC) can play a decisive role: the analysis code should be scalable so that multiple graphical processing units (GPUs) and compute nodes can be efficiently utilized to reduce runtime. HPC systems provide parallel file systems with high input/output (I/O) throughput, enabling efficient data reading and writing. Both computing and data infrastructures support optimization techniques that can further enhance performance. Finally, HPC environments provide authentication and authorization mechanisms to manage access rights to resources, thereby facilitating secure data sharing and collaboration. Overall, this work illustrates how combining graph-based modeling with scalable HPC infrastructure could potentially enable the analysis of complex patient networks, paving the way for large-scale, bias-aware clustering approaches in real-world clinical data.

## Data Availability

All data produced in the present study are available upon reasonable request to the authors

## Ethical approval

This study was approved by the Ethics Committee at the Baden-Württemberg State Medical Association (Ethik-Kommission der Landesärztekammer Baden-Württemberg), with approval number F-2024-105. The study was performed in compliance with the World Medical Association Declaration of Helsinki on Ethical Principles for Medical Research Involving Human Subjects and research regulations of the country.

